# ANTIMICROBIAL SUSCEPTIBILITY PATTERNS OF BACTERIAL PATHOGENS IN ACUTE EXACERBATIONS OF CHRONIC OBSTRUCTIVE PULMONARY DISEASE IN PATIENTS AT A TERTIARY CARE FACILITY

**DOI:** 10.1101/2024.11.19.24317537

**Authors:** Susma Dahal, Renuka Thapa, Puja Gurung, Abinas Chaudhary, Dinesh Dhakal, Upendra Thapa Shrestha, Raina Chaudhary, Amrit Acharya

## Abstract

**Objectives:** The study was aimed to determine the predominant bacteria causing Acute Exacerbations of Chronic Obstructive Pulmonary Disease (AECOPD) infection in patients and their antibiotics sensitivity pattern in tertiary care setup.

**Methods:** This descriptive Cross-sectional study was conducted from September 27^th^ 2023 to December 26^th^ 2023 at Shree Birendra Hospital, chhauni, Kathmandu. Sputum samples that were received in the Microbiology laboratory from in patients for routine diagnosis were included in the study. The samples that were received was subjected to gram staining to assess the quality of the sputum sample, and those samples with good quality (mucoid and muco-purulent) was inoculated onto Blood agar, Chocolate agar and Mac Conkey agar for the isolation of the pathogens, and the media was incubated at 37 degrees Celsius overnight. Culture isolates were identified by standard technique(Cowan and Steel’s Manual for the Identification of Medical Bacteria, 1993) and Kirby-Bauer method was used to test antibiotic sensitivity of the pathogenic organisms following protocol of (Clinical and Laboratory Standards Institute, 2018) guidelines.

**Results:** Out of 273 sputum samples, 42/273 (15.4%) showed growth. Five different bacterial species were identified. Among the isolates, *Acinetobacter* spp. was the most common pathogen 21(44.7%) followed by *Pseudomonas aeruginosa* 11(23.4%), *Klebsiella pneumoniae* 10(21.3%), *Escherichia coli* 4(8.5%) and *Citrobacter freundii 1(*2.1%). Highest number of AECOPD cases were observed in female 157(57.9%) with 25(15.9%) positivity and highest number of organisms were isolated in age group 56-70(17) and least in age group 40-55(3).

All the *Acinetobacter* spp. (n=21) isolates were resistant to all tested medicine. Almost 90.90% *Pseudomonas aeruginosa* (n=11) were sensitive to Gentamycin and 81.82% to Meropenem, 70% *Klebsiella pneumoniae* (n=10) were sensitive to Gentamycin, 75% *Escherichia coli* (n=4) and 100 % *Citrobacter freundii* (n=1) to Amikacin. 100% (n=47) isolates were resistant to antibiotic penicillin and 100% *Escherichia coli and Citrobacter freundii were resistance to* Ceftriazone and Cefepime while 100% (n=47) were found to be multi-drug resistant.

**Conclusion:** *Acinetobacter* spp *P. aeruginosa, Klebsiella pneumoniae*, and *Escherichia coli* were the most common bacterial isolates in the current investigation, which revealed a 15.4 % of culture positive. There is a high rate of MDR pattern in all isolated isolates. To further enhance treatment quality and prevent antibiotic resistance, regular surveillance of the etiologies of AECOPD and their pattern of antimicrobial susceptibility is crucial.

## Introduction

Physiologically COPD is defined by the presence of irreversible or partially reversible airway obstruction in patients with chronic bronchitis and emphysema (“Standards for the Diagnosis and Care of Patients with Chronic Obstructive Pulmonary Disease. American Thoracic Society,” 1995). Exacerbations of chronic obstructive pulmonary disease (AECOPD) is the prevalent disease in the world (Pauwels et al., 2001) (Chronic obstructive pulmonary disease (COPD) is a disease, the incidence of which continues to increase(Mannino et al., 2006) .It is the leading cause of death worldwide, with a rising mortality rate, and it is expected that this will be the fifth leading condition among conditions with a high socioeconomic burden to Society(Hurd, 2000) .

Exacerbations are common for many patients with chronic obstructive pulmonary disease (COPD) and contribute greatly to an increase in morbidity, frequent emergency department (ED) visits, hospital admissions, and increased healthcare costs (Bourbeau et al., 2003; Connors et al., 1996; Donaldson et al., 2002; Hurd, 2000) Evidence suggests that patients with recurrent acute exacerbations of COPD (AECOPD) have a faster decline in lung function (Connors et al., 1996; Donaldson et al., 2002), possibly due to increased rate of airway inflammation (Kanner et al., 2001). The health impacts of AECOPD infection can be severe and may include Worsening of COPD symptoms, increased risk of complications, reduced quality of life, increased healthcare costs. AECOPD infection can have a significant impact on a person’s health and well-being. It is important to take steps to prevent and manage the condition, such as quitting smoking, getting regular check-ups with a health care provider, and following a treatment plan (Donaldson et al., 2006; Miravitlles et al., 2004; Seemungal et al., 1998).

Chronic obstructive pulmonary disease (COPD) is the third leading cause of death worldwide, causing 3.23 million deaths in 2019.Nearly 90% of COPD deaths in those under 70 years of age occur in low- and middle-income countries (LMIC) (Spencer & Jones, 2003). Most patients admitted for treatment of COPD were women (60%) and from higher ethnic groups (having a comparative advantage in terms of social and economic status), with a greater prevalence among those aged 60-69 years (37% of overall cases). The incidence of COPD increased in consecutive years, with the highest load during the winter month. COPD is a significant health burden in Nepal, primarily due to high rates of smoking and exposure to indoor and outdoor air pollution. The prevalence of COPD in Nepal varies depending on the region, with higher rates reported in rural areas compared to urban areas. AECOPD is a common complication of COPD, characterized by exacerbation of respiratory symptoms such as cough, sputum production, dyspnea, and wheezing. AECOPD is a leading cause of hospitalization and mortality among COPD patients worldwide. In Nepal, there is a limited amount of epidemiological data regarding AECOPD. However, a study conducted in 2017 among COPD patients in Kathmandu Valley reported that around 80% of patients experienced at least one episode of AECOPD in the previous year, with a mean annual exacerbation rate of around 2.3 episodes per patient. The study also found that exposure to biomass fuel and a history of hospitalization for AECOPD was significant predictors of future exacerbation (Adhikari et al., 2021; Karchmer, 2004) Sample obtained by bronchoscopy studies, analysis of human immune response with appropriate immunoassay, and antibiotic trials display approximately half of the exacerbation are caused by bacteria (Sethi & Murphy, 2001). The predominant microorganisms isolated from the sputum sample are *Haemophilus influenza, Streptococcus pneumonia, Pseudomonas aeruginosa, Enterobacteriaceae, Staphylococcus aureus, and Moraxella catarrhalis* (Vestbo et al., 2013).

Both the dominance of morbidity and mortality from COPD have been rising globally, this is due to the increasing antimicrobial resistance (AMR). High rates of occurrence and rising antimicrobial resistance among microorganisms causing COPD greatly to increase the economic and social burden of these diseases. The lack of appropriate therapeutics to encounter the resistant pathogen has enhanced the urge for the evolution of either new antibiotics or different therapeutic combinations. Hence this study’s purpose is to isolate predominant microorganisms causing COPD. This study will fill the gap of investigation of COPD and will generate knowledge about multidrug resistance microorganism, their antimicrobial susceptibility pattern which help to study pathogenesis of microorganism, disease pattern to cure disease or to decrease the burden from the disease. This investigation will be the knowledge to develop antibiotics or combination of antibiotics to cure disease in future.

## Methods and Methodology

### Methodology

#### 1 Study design

Descriptive Cross-sectional study was performed.

#### 2 Sampling method and sample size

Non-probability consecutive sampling technique was used for patients’ selection. The previous sub-national studies conducted in Nepal have reported the prevalence of COPD were 23%.(Adhikari et al., 2018, 2020) . So, sample size of around 273 AECOPD patients required for this study.

#### 3 Study sites

The study were carried out at Shree Birendra Hospital, Chhauni and Laboratory analysis was conducted at Microbiology Laboratory, Shree Birendra Hospital, Chhauni, Kathmandu.

#### 4 Study populations

The entire hospitalized AECOPD patient in the medical ward, ICU, and surgical ward of Shree Birendra Hospital of age (40+), any sex within the study duration were considered as population.

#### 5 Study period and ethical considerations

The study was conducted from August 2023-October2023, after the proposal was approved by Institutional review committee (IRC), Nepalese Army Institute of Health Sciences (NAIHS).

#### 6 Sampling Units

Patients with AECOPD admitted in the intensive care unit were included.

#### 7 Data Collection/ techniques

Inpatient’s Sputum samples that received in the Microbiology laboratory for routine diagnosis were included in the study. The samples were processed according to the departmental standard operating procedures. In brief, the samples that had received were subjected to gram staining to assess the quality of the sputum sample, and those samples with good quality (mucoid and muco-purulent) were inoculated onto Blood agar, Chocolate agar and Mac Conkey agar for the isolation of the pathogens, and the media were incubated at 37 degrees Celsius overnight. The growth was further identified and antibiotic susceptibility testing of the isolates was determined following the protocol of CLSI, (2020) guidelines.

##### 7.1 Data Collection Tools

All diagnosed patients with AECOPD by the concerned clinician depending upon the presence of two of the following symptoms having acute exacerbation with increased dyspnea, increased sputum volume, increased sputum purulence, and adequate sputum sample based on gram staining.

Socio-demographic characteristics and clinical of the patient were collected by face-to-face administered semi-structured questionnaires using study variables. Expectorate d sputum specimens were collected in a sterile sputum cup (falcon tube) by standard collection procedures, then collected samples were proceed for microbiological test.

##### 7.2 Specimen Culture

Samples were accepted after microscopic evaluation and washing procedures, sputum was inoculated directly onto blood agar, Mac Conkey agar, and chocolate agar for bacterial isolation. The chocolate agar plate was incubated in an incubator (5–10% CO2) at 37°C for 24–48 hours, while blood agar and Mac Conkey agar, were incubated in an aerobic atmosphere at 37°C for 24 hours. The suspicious colony were subculture on suitable solid culture media for purification, thereafter further procedures were processed or preserved on appropriate media and stored in a refrigerator (4°C) for subsequent analysis as necessary. Isolated micro-organisms were considered significant and accepted as causative pathogens only if they reached a count of 10^6^ CFU/ml according to (Cheesbrough, 2006).

##### 7.3 Identification of Isolated Organisms

The identification of isolates was accomplished using the standard microbiological techniques, which involved morphological colony studies, Gram staining, and a battery of biochemical tests like Catalase test, Oxidase test, Indole, Citrate utilization test, Urease production, Hydrogen sulfide production, Sugar fermentation test, Coagulase test, and Optochin sensitivity tests.

##### 7.4 Antibiotic susceptibility testing and multidrug-resistant

The antimicrobial susceptibility pattern of isolates was done using the Kirby Bauer disk diffusion method on Mueller Hinton agar using Penicillin, Cefoxitin, Clindamycin, and Erythromycin for Gram-positive bacteria only. Tetracycline, Ampicillin, Chloramphenicol, Gentamycin, Ciprofloxacin, Trimethoprim-sulphamethoxazole, Ceftriaxone, Amoxicillin-clavulanic acid, and Cefepime, for both Gram-positives and Gram-negatives but Amikacin, Tobramycin, and Meropenem for Gram-negative bacteria only according to (CLSI, 2020). Similarly, the isolates that were resistant to 3 or more different classes of antibiotics were considered multidrug-resistant strains (Magiorakos et al., 2012) .

#### 8. Potential biases

All patients with known cases of tuberculosis, pneumonia, and asthma with evidence of either clinically or radiologically and patients, who were unable to participate, excluded from the study.

#### 9. Quality Control

Quality control must be maintained in order to get reliable microbiological results. Utilizing the guidelines provided by the (Clinical and Laboratory Standards Institute, 2018), quality control was carried out throughout this investigation in monitoring of the laboratory equipment’s, reagents and media, quality control during isolation and identification of pathogenic bacteria.

## RESULT

### Growth Pattern of the specimen

Out of 273 sputum sample, 42 (15.4%) showed growth and 231(84.6%) was found to be no growth with upper respiratory tract normal flora. The results have shown in figure 1.

**Figure 1:**
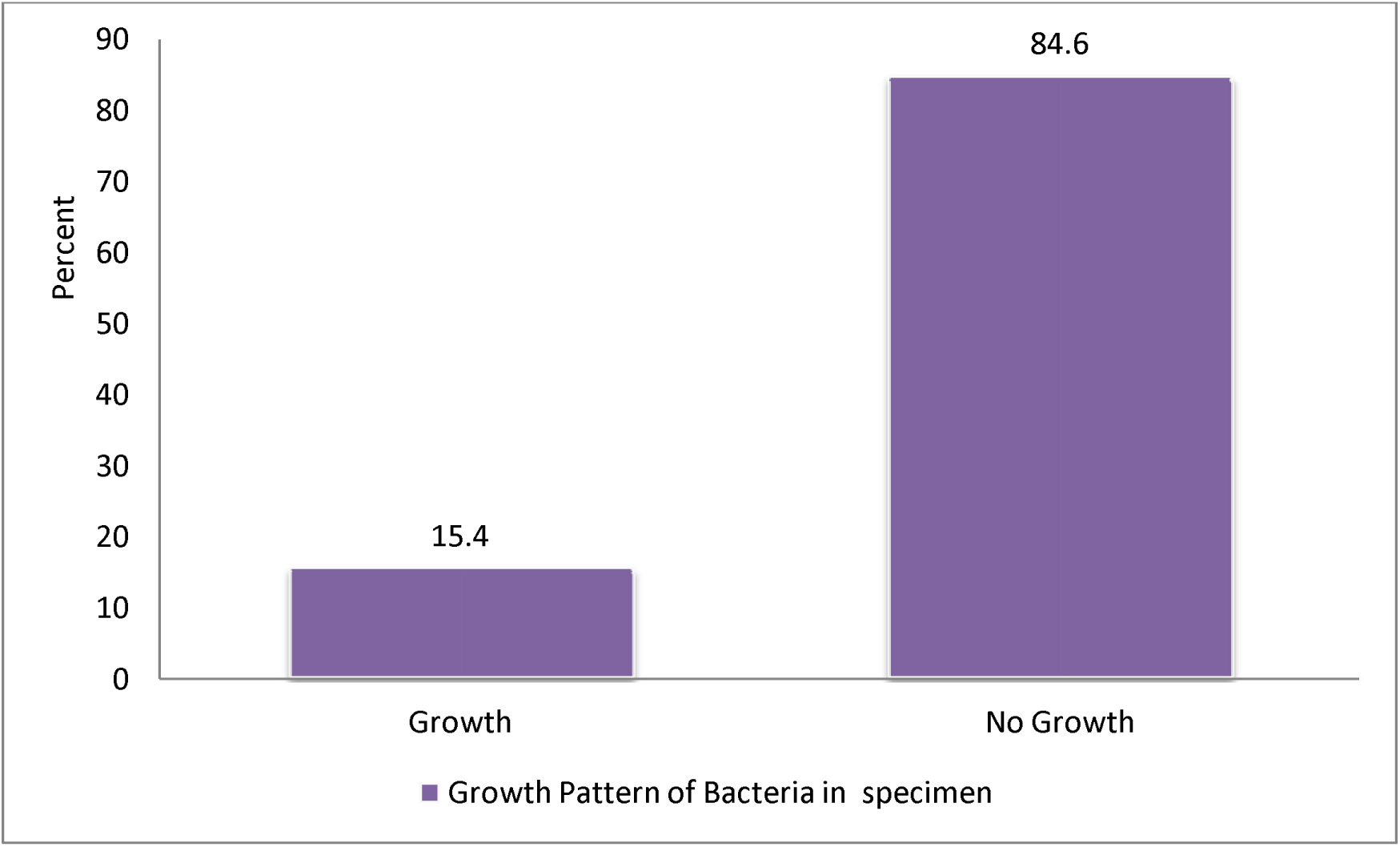
Growth Pattern of Bacteria in specimen **Pattern of MDR among bacterial isolates** All the 47 bacterial isolates were observed multi-drug resistance.

### Age-wise distribution of AECOPD

Among the 273 AECOPD patients, AEOPD was observed high in age group 71-85 with 132(48.35%) while least in age group 40-55 with 17(6.23%). Highest number of organism is isolated in age group 56-70(17) and least in age group 40-55(3) (table 1).

**Table 1:** Age- wise distribution of AECOPD.

| S.N | Age | Total | Positivity (%) | MDR |
| --- | --- | --- | --- | --- |
| 1. | Below 40 | 0 | 0(0) | 0(0) |
| 2. | 40-55 | 17 | 3 (17.65) | 3(100) |
| 3. | 56-70 | 103 | 17( 16.50) | 17(100) |
| 4. | 71-85 | 132 | 17(12.88) | 17(100) |
| **5** | Above 85 | 21 | 5(23.81) | 5(100) |
| **Total** |  | **273** | **42(70.84)** | **42(100)** |

### Gender-wise distribution of AECOPD and bacterial isolates

Among 273 samples, highest number of AECOPD cases were observed in female 157(57.9%) with 25(15.9%) positivity. In male, 116(42.5%) cases were observed with 17(6.2%) positivity during study time. (Table 2)

**Table 2:** Gender-wise distribution of AECOPD and bacterial isolates.

| **S.N** | **Gender** | **Frequency(%)**<br>**n=273** | **Positivity(%)**<br>**n=42** | **p- Value** |
| --- | --- | --- | --- | --- |
| **1.** | Female | 157(57.5) | 25(9.2) | P= 0.906 |
| **2.** | Male | 116(42.5) | 17(6.2) |  |
|  | **Total** | **273(100)** | **42(15.4)** |  |

**Table 3:**
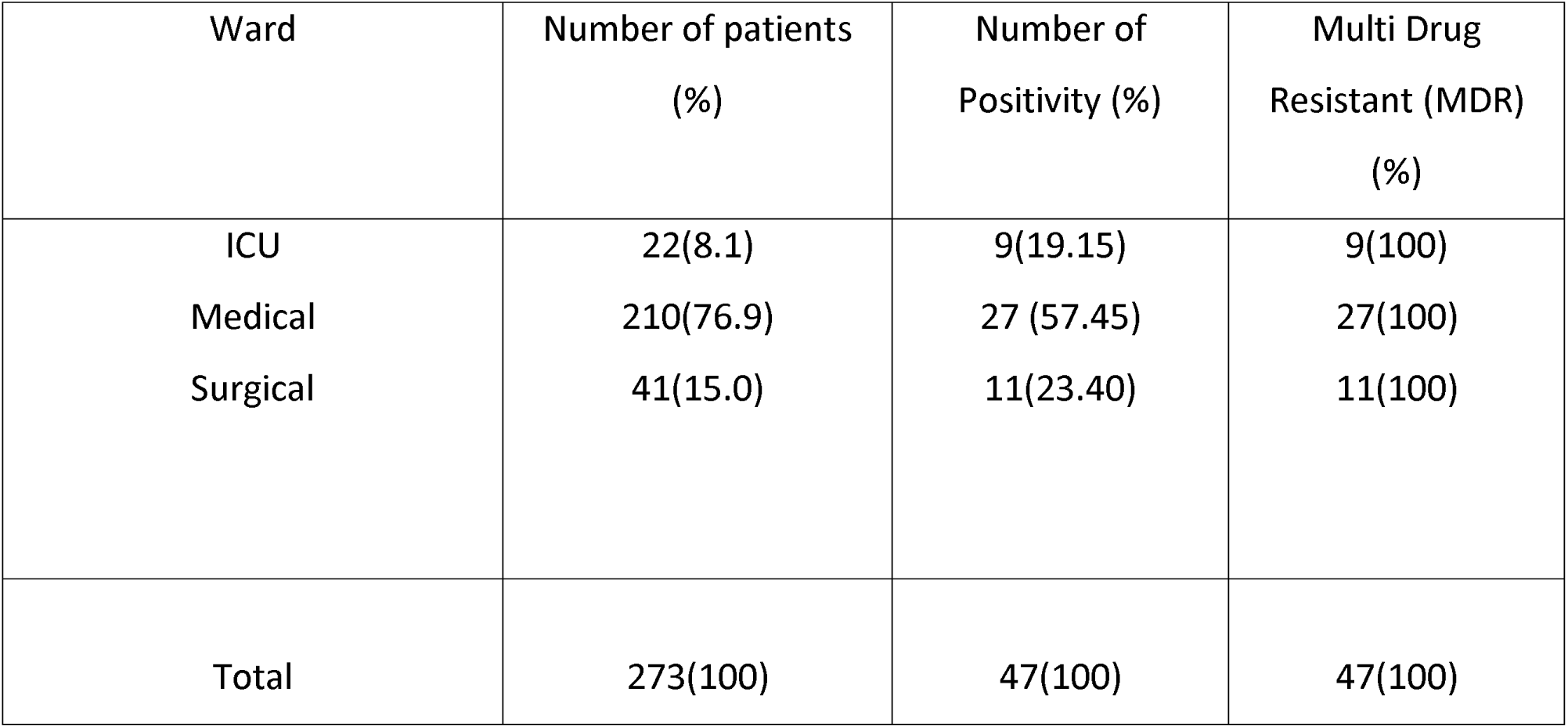
Ward-wise Multi-Drug Resistant (MDR) pattern of Bacterial isolates.

| Ward | Number of patients<br>(%) | Number of<br>Positivity (%) | Multi Drug<br>Resistant (MDR)<br>(%) |
| --- | --- | --- | --- |
| ICU | 22(8.1) | 9(19.15) | 9(100) |
| Medical | 210(76.9) | 27 (57.45) | 27(100) |
| Surgical | 41(15.0) | 11(23.40) | 11(100) |
| Total | 273(100) | 47(100) | 47(100) |

**Table 4:**
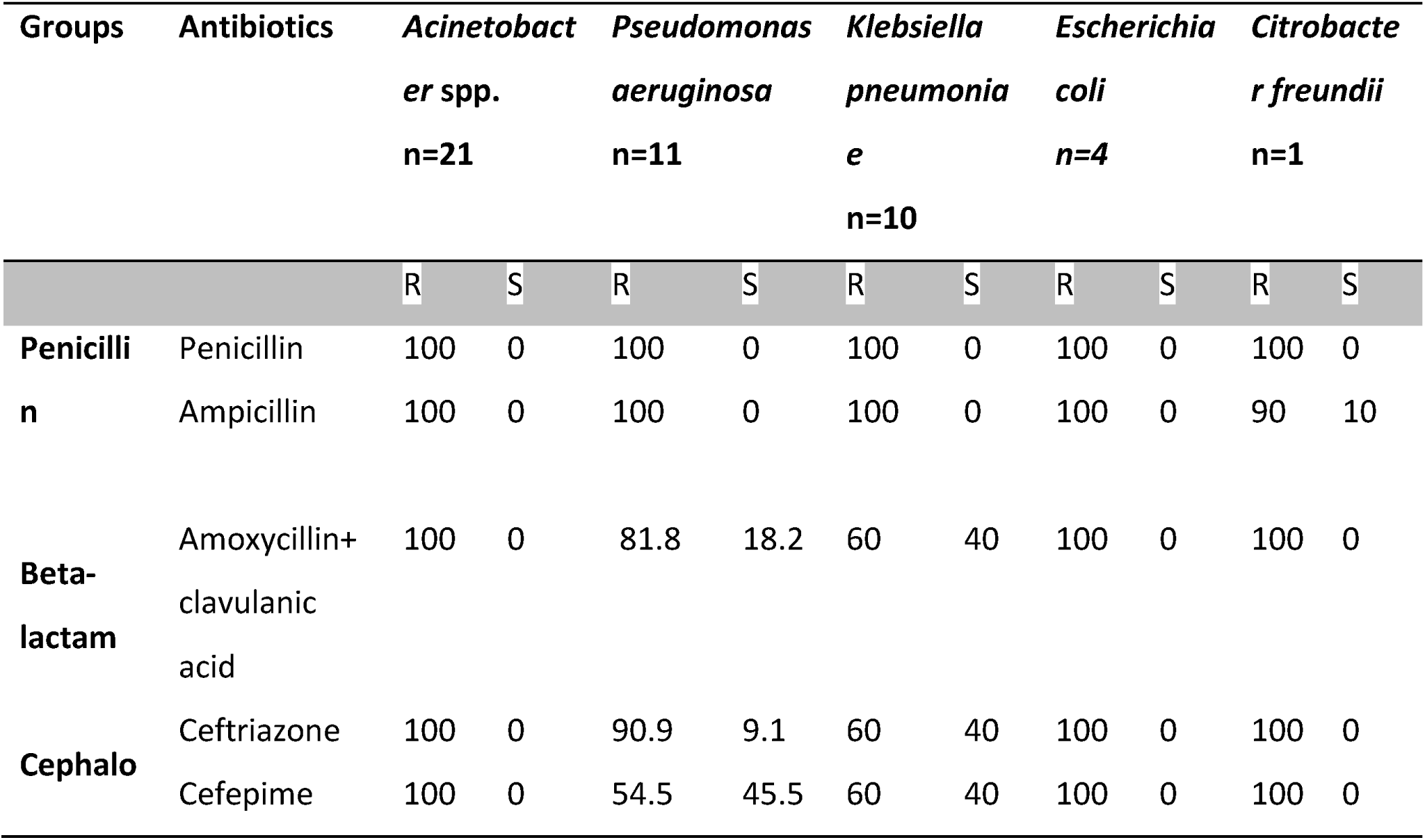
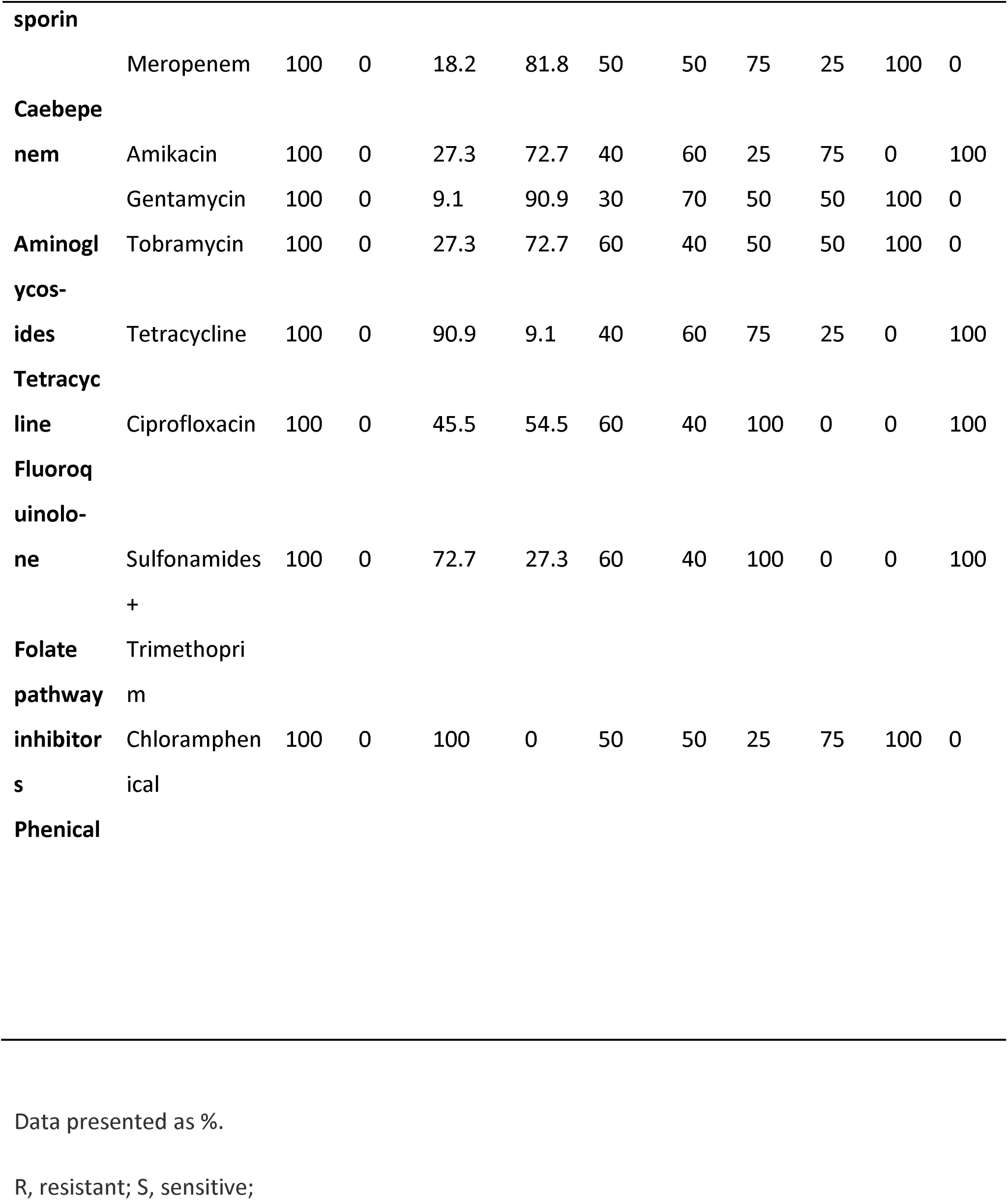
Antibiotic susceptibility of the most commonly isolated. Amino glycosides antibiotic group showed more effective against all bacterial isolate than other antibiotics group.

**Table 5:** Pattern of MDR among bacterial isolates.

| S.N | Organisms | Total isolates | MDR | p- value |
| --- | --- | --- | --- | --- |
|  |  |  | Number (%) |  |
| 1. | *Acinetobacter spp.* | 21 | 21(100) | P= 0.00001 |
| 2. | *Pseudomonas aeruginosa* | 11 | 11(100) |  |
| 3. | *Klebsiella pneumoniae* | 10 | 10(100) |  |
| 4. | *Escherichia coli* | 4 | 4(100) |  |
| 5. | *Citrobacter freundii* | 1 | 1(100) |  |
| Total |  | 47 | 47(100) |  |

**Table 6:** Phenotype resistance of isolated bacteria.

| S.N | Organisms | Phenotype of resistance | Total number |
| --- | --- | --- | --- |
| 1 | *Acinetobacter spp.* | P,AMP,AUG,CRO,CPM,MEM,AK,GN, TM,T,CIP,COT,C | 13 |
| 2 | *Pseudomonas* | P,AMP,AUG,CRO,T,COT,C | 7 |
| 3 | *aeruginosa* | P,AMP,AUG,CRO,CPM,TM,CIP,COT | 8 |
| 4 | *Klebsiella pneumoniae* | P,AMP,AUG,CRO,CPM,MEM,T,CIP, | 9 |
|  | *Escherichia coli* | COT |  |
| 5 |  | P,AMP,AUG,CRO,CPM,MEM,GN,TN,C | 9 |
|  | *Citrobacter freundii* |  |  |

### Ward-wise distribution of AECOPD and bacterial isolates

Among 273 cases, 210(76.9%) samples from medical ward, 41(15.0%) from surgical ward and 22(8.1%) from ICU were observed as shown in (Figure 2).

**Figure 2:**
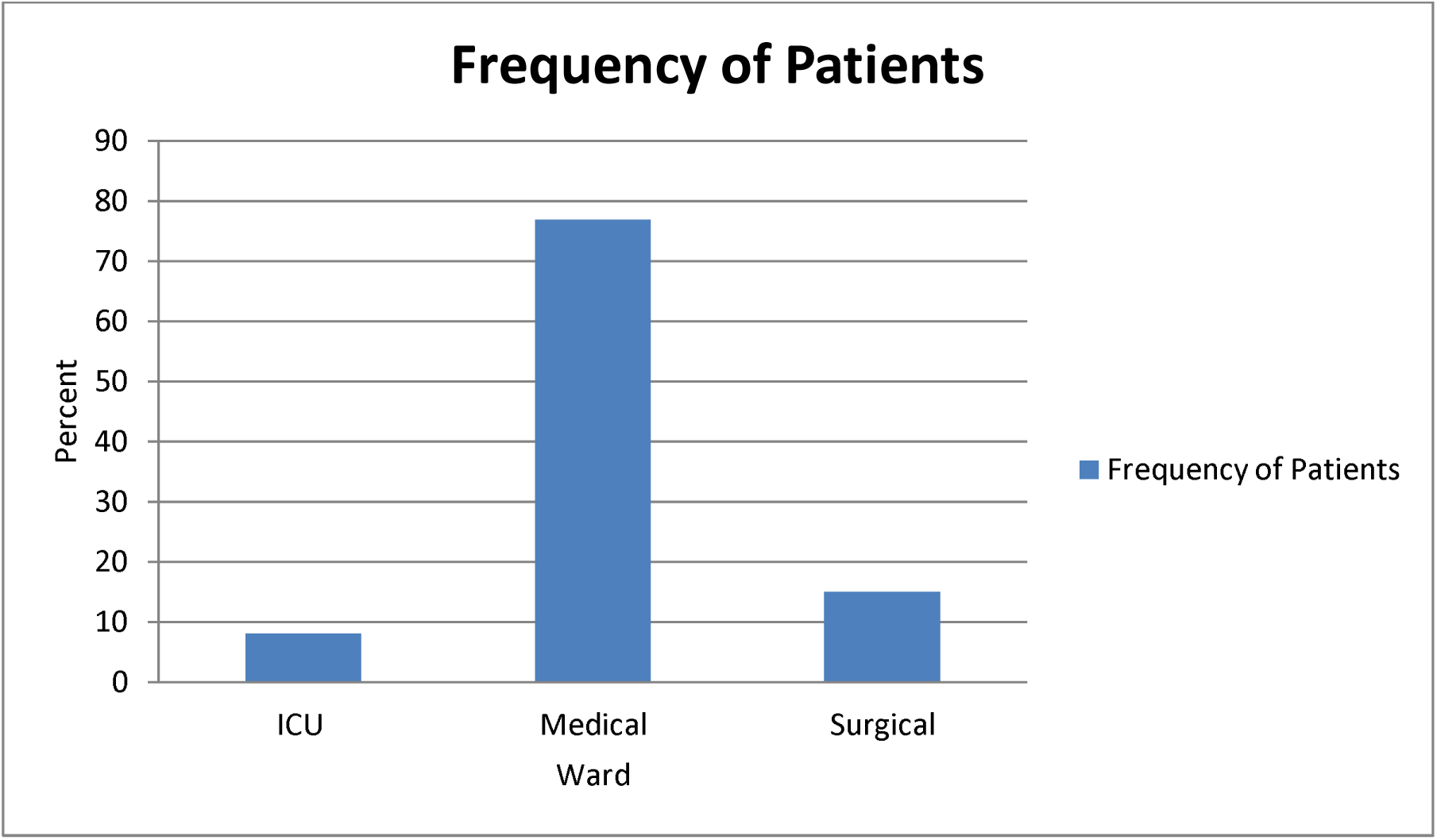
Ward-wise distribution of bacterial isolates from COPD Patients

### Pattern of bacterial isolates in culture results

Among 42 growths, all organisms were observed Gram negative bacteria. *Acinetobacter* spp. was observed in highest number 21(44.7%) and *Citrobacter freundii* in least cases 1(2.1%).(Figure 3)

**Figure 3:**
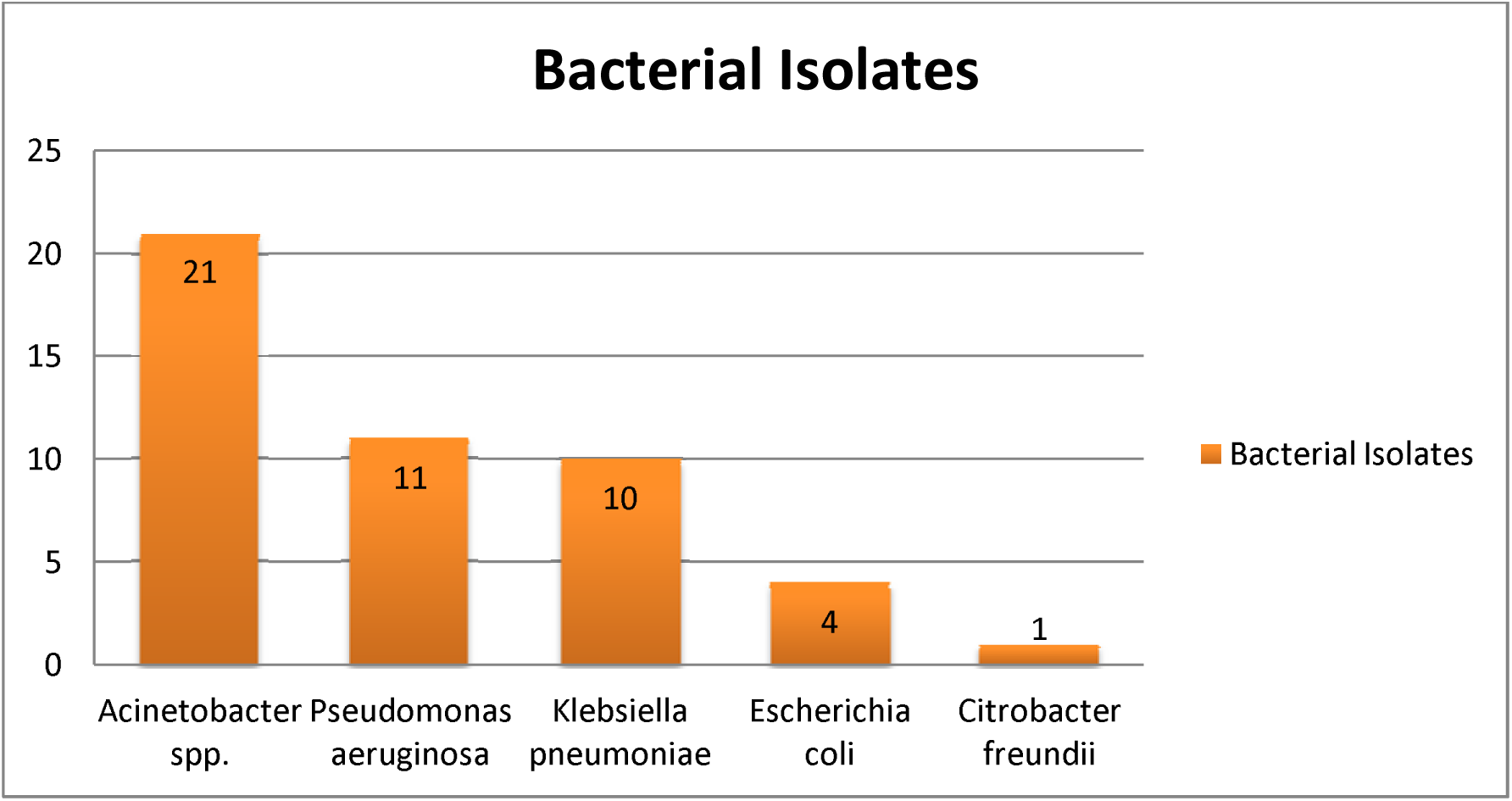
Pattern of bacterial isolates in culture result

### Antibiotic susceptibility

Amino glycosides antibiotic group showed more effective against all bacterial isolate than other antibiotics group. All the *Acinetobacter* spp. (n=21) isolates were resistant to all tested medicine. Almost 78.77% *Pseudomonas aeruginosa* (n=11) were sensitive to Aminoglycoside, 70% *Klebsiella pneumoniae* (n=10) were sensitive to Gentamycin, 75% *Escherichia coli* (n=4) and 100 % *Citrobacter freundii* (n=1) to Amikacin. 100% (n=47) isolates were resistant to antibiotic penicillin and 100% *Escherichia coli and Citrobacter freundiiwere resistance to* Ceftriazone and Cefepime while 100% (n=42) were found to be multi-drug resistant.

### Discussion

The acute exacerbation of chronic obstructive pulmonary disease has been believed to be primarily caused by bacterial-induced respiratory infections. About half of COPD exacerbations are linked to bacteria isolated from the lower respiratory tract(Beasley et al., 2012; Sethi, 2000). Therefore, this study provides insight on treatment of pathogenic bacteria located in lower respiratory tract. In this study, overall Two hundred Seventy three Patients with AECOPD were included, Growth of pathogenic organisms were seen in 42(15.4%) which was very similar with the study conducted in India by Ullah et al., (2022) with 14.3% growth rate but difference result was seen in the study of Saxena et al., (2016) with (42%).This variance in the positivity of respiratory diseases between groups and nations, dependent on a difference in location, climate, and socioeconomic situations, as well as it may be attributed to antibiotic abuse (Ferrera et al., 2021; Zhou et al., 2024).

In this study, maximum AECOPD was observed in the age group 71-85 with 48.35%. Previous study by Raherison & Girodet, (2009), those over 65 years of age have a by five times higher chance of getting COPD than those under 40 years of age. Also in the study by Anecchino et al.,(2007), examined a cohort of 126,283 COPD patients (≥45 years) found that the prevalence of the disease was 3.6% and that it rose sharply with age (1.9% in those 45–64 years old, 4.8% in those 65–74 years old, 6.8% in those 75–84 years old, and 5.6% in those ≥ 85 years old).

And high growth positivity was found in age group 56-70 with 16.5% positivity rate which was comparable with study conducted by Lal et al., (2017) in India where higher positivity rate was 40%, from age group 55-65years. Lower microbial rate in higher age may be due to the influence of potentially modifiable factors, such as preterm birth, smoke exposure, recurrent pulmonary infections, and persistent asthma during childhood, may be reflected in the lung function trajectories from birth or childhood to early adulthood, as identified by multiple birth-cohort studies. These factors could be the focus of interventions to maximize lung growth and lower the risk of COPD and infection in later life(Belgrave et al., 2018; Bui et al., 2018; Casas et al., 2018; den Dekker et al., 2018).

In our study, the prevalence of Aecopd was higher among the females compared to male. And also bacterial positivity was found higher in the female (9.2%).This is similar to a study report by Elfeky et al., (2016) exposure to indoor air pollution was more common in females than nonsmokers. Regarding gender differences, study by Bouquet et al., (2020) have shown that women with COPD may experience more frequent and severe exacerbations compared to men. This could be due to differences in immune response, hormonal influences and variation in lung microbiota composition.

In the current study, all the bacteria isolates were reported Gram-negative bacteria. However, in the observation by Arafa, (2021); Elfeky et al., (2016) was shown that 80% of the isolates were Gram-positive. But different result can observed in the study of Bouquet et al., (2020), where *Haemophilus and Moraxella* were predominant in acute exacerbation samples. There may be a significant correlation between the type of study participants and the disparity in the prevalence of bacterial infections. This investigation focused on hospitalized patients who had been hospitalized because their COPD significantly worsened. These patients are more likely to be infected by Gram-negative bacteria, while community-based outpatients were more likely to be mostly Gram-positive (Kuwal et al., 2018; Saxena et al., 2016). In this study we were able to isolate bacterial colony from single and mixed colony.

When discussing particular bacterial isolates, *Acinetobacter* spp. (44.7%), was the most common bacteria isolated in our study,followed by *P. aeruginosa(23.4%).*Prior research conducted by Millares et al., (2014) demonstrated that patients with more severe COPD have a higher frequency of chronic *P. aeruginosa* colonization.While other studies have implicated K. *pneumoniae* and *P. aeruginosa* were predominance bacteria responsible for Aecopd (Kuwal et al., 2018; Madhavi et al., 2017). The variation in culture positive and distribution of microorganisms might be as a result of differing collecting techniques, transportation time, the number of organisms presents in the sample, and the nature of the sputum (Saxena et al., 2016; Sethi & Murphy, 2008) . Furthermore, studies focused specifically on microorganisms from the western population in AECOPD patients found that *S. pneumoniae, H. influenzae, or S. aureus* were the most common, followed by Gram-negative bacteria (Sethi & Murphy, 2001; Simpson et al., 2016).

For the Antibiotic Susceptibility pattern *Acinetobacter* Spp. was resistant to almost all antimicrobial agents that were used. Increasing resistance of *Acinetobacter* Spp. has been reported globally in the last decade (Ullah et al., 2022; Wang et al., 2010) .Infection with *Acinetobacter* spp. in COPD patient are one of the major concerns to treat because of their intrinsic and acquired resistance.

Overall, antibiotic susceptibility test showed that Gram negative bacteria were most effective to Aminoglycosides group of antibiotics. This study showed that all (100%) of the bacterial isolates were MDR. This was higher than study conducted by Ullah et al., (2022,) in south India (57.6%) MDR rate and very similar to study conducted in Southwest Ethopia (93.8%) MDR rate by Mussema et al., (2022). This could be because of the frequent prescriptions for these antibiotics, which lead to their regular usage and the emergence of drug resistance or inappropriate antimicrobial use (Gillespie et al., 2021). Almost 78.77% *Pseudomonas aeruginosa* (n=11) were sensitive to Aminoglycoside, 70% *Klebsiella pneumoniae* (n=10) were sensitive to Gentamycin, 75% *Escherichia coli* (n=4) and 100 % *Citrobacter freundii* (n=1) to Amikacin. 100% (n=47) isolates were resistant to antibiotic penicillin and 100% *Escherichia coli and Citrobacter freundiiwere resistance to* Ceftriazone and Cefepime while 100% (n=42) were found to be multi- drug resistant. The World Health Organization officially lists antibiotic resistance as one of the main risks to public health worldwide(WHO, 2015). We are currently dealing with a global crisis that has multiple troubling aspects: the majority of clinical isolates of bacteria have resistance mechanisms, new antibiotics are no longer being discovered, and recurrent infections brought on by persistent bacteria are making it more difficult to treat infections successfully.

These data help clinicians informed of which antibiotics are appropriate for use. This study presented information on AECOPD-related infections that may help guide antibiotic choices in the therapy of AECOPD in Nepal. At last, our findings have many similarities and contrasts with previous research. Continued surveillance, particularly based on local data, is clearly required to understand the issues of antimicrobial resistance and prevent its spread.

## Data Availability

All data produced in the present study are available upon reasonable request to the authors.
If all data are included in the manuscript, you can alternatively use:
All data produced in the present work are contained in the manuscript.
Choose the one that best fits your data-sharing approach.

Https://www.nepjol.info

## Acknowledgements

We express our sincere thanks the staff and management of Shree Birendra Hospital for their support and cooperation. Special appreciation goes to the microbiology laboratory team, for their invaluable assistance and guidance. We also thank administrative staff and the patients who participated in our study.

We would like to thanks Dal Prasad Pun, principal, Colonel Arjun Adhikari, Head of management, Mahendra Bam, Vice Principal of Sainik Awasiya Mahavidhyalaya for their invaluable support and guidance. We express our gratitude toward Department of Microbiology and Subhash Kumar Verma, B.Sc. Co- coordinator, Sainik Awasiya Mahavidhyalaya for support to carry out this research.

